# Causative Organisms, Antibiotic Sensitivity Patterns and Risk Factors Associated with Neonatal Sepsis at Moi Teaching and Referral Hospital, Kenya

**DOI:** 10.1101/2020.12.02.20242594

**Authors:** Benard Mageto Ateka, Julia Songok, Winstone Nyandiko

## Abstract

**Background:** Neonatal bacterial infections have been associated with rising antimicrobial resistance levels. This has led to increasing neonatal morbidity and mortality in poorly resourced health facilities located in low income countries. Local studies on neonatal antibiotic sensitivity patterns and its associated risk factors could inform empirical antibiotic therapy and hospital infection control strategies.

**Purpose:** The study aimed at determining the causative organisms, antibiotic sensitivity patterns and risk factors associated with neonatal sepsis at a tertiary teaching hospital in Western Kenya.

**Materials and Methods:** Cross-sectional study among neonates suspected to have sepsis and undergoing treatment at Moi Teaching and Referral Hospital (MTRH’s) newborn unit (NBU) between September 2017 and July 2018. Blood culture tests isolated bacteria and determined their antimicrobial sensitivity. Neonatal and maternal characteristics were obtained through medical chart reviews. Descriptive statistics, Pearson chi-square test of association and odds ratios were adopted.

**Results:** The study enrolled 141 neonates, majority (57.4%) of whom were female. The median gestational age and birth weight were 37 (IQR: 22-45) weeks and 2400g (IQR: 800 - 4700) respectively. Of the 151 bacterial isolates identified, 46.4% were *Klebsiella spp*. followed by *Coagulate negative staphylococcus* (CoNS) at 27.8%. *Klebsiella spp*. was sensitive to meropenem, amikacin and cefepime but resistant to ceftriaxone, gentamycin and cefotaxime. However, CoNS was sensitive to vancomycin and penicillin. Both the neonatal and maternal risk factors assessed were not associated with neonatal sepsis.

**Conclusion:** The main bacterial causes of neonatal sepsis were *Klebsiella spp*. and CoNS which were both sensitivity to meropenem and amikacin.

## INTRODUCTION

Neonatal sepsis is a clinical syndrome in an infant up to 28 days of age, which manifests by systemic signs of infection [1]. It can be confirmed by isolating the causative pathogen(s) from the blood stream, as it is a major cause of neonatal morbidity and mortality globally [2]. Approximately 2 million of the 30 million neonates who contract infections annually around the globe succumb to neonatal sepsis [3]. Although the neonatal mortality rate in the developed economies stand at 0.69 deaths per 1000 live births; higher rates of 0.76 deaths per 1000 live births have been reported in the low and middle income countries [3]. The global incidence rate of neonatal sepsis is at 22 per 1000 live births with an estimated mortality of 11%-19% [4]. The mortality rate due to neonatal sepsis in Africa has been estimated at 12% in Nigeria [5] and 28% in Kenya [6].

Neonatal sepsis is commonly caused by bacteria that can be classified based on gram staining patterns as either gram positive or gram negative. In countries with developing economies, gram positive organisms such as coagulase negative *Staphylococci* (CoNS), *Enterococcus faecalis*, methicillin-resistant and methicillin-susceptible *Staphylococcus aureus*-MRSA/MSSA and *Streptococcus pneumoniae* are the most predominant causative organism [7]. On the other hand, gram negative bacteria such as *Klebsiella pneumoniae, Serratia marcescens, Acinetobacter baumannii and Escherichia coli* have been associated with neonatal sepsis in high-income countries [8]. Neonatal sepsis [3] can be classified further based on the time of onset of symptoms as either early onset (≤72 hours of life) or late onset (>72 hours of life) [2,9–11]. Early onset neonatal sepsis (EONS) is associated with causative factors [12] of maternal origin (vertical transmission); while late onset neonatal sepsis (LONS) could be either community acquired or nosocomial among hospitalized neonates [11,13,14]. In the United Kingdom, early onset neonatal sepsis associated with gram negative bacteria has been reported to be susceptible to a combination of penicillin and gentamicin (94%), amoxicillin and cefotaxime (100%), amoxicillin and other penicillin (98%) and cefotaxime monotherapy at 96% [15]. However, majority of the gram-positive bacteria resisted these treatment combinations. Frequent use of third generation cephalosporins drive the development of resistance bacterial pathogens in neonatal intensive care units and could lead to an emergence of extended spectrum beta lactamase producing strains. Due to limited data in Western Kenya, the antibiotic sensitivity patterns of these bacteria are not well known.

Most early onset neonatal sepsis are associated with both maternal and neonatal factors [16]. Maternal factors include age of the mother, level of education, intrapartum pyrexia, parity, attendance of antenatal clinic and presence of urinary tract infection in pregnancy [2,17]. Neonatal risk factors include: APGAR (appearance, pulse, grimace, activity and respiration) score, mode of delivery and gestational maturity [13,18]. There is need to determine the causative organisms of neonatal sepsis in clinical settings to inform localized infection control strategies, improve on patient care and outcome. Profiling of the sensitivity patterns of the specific pathogens to the commonly used antibiotics inform empirical therapy selection. This study therefore aimed at determining the causative organisms, antibiotic sensitivity patterns and risk factors associated with neonatal sepsis.

## MATERIALS AND METHODS

This was a cross-sectional descriptive study conducted at the newborn unit (NBU) of Moi Teaching and Referral Hospital (MTRH). The facility which is the second largest referral hospital in Kenya and is in the Western part of the country in Uasin Gishu. The study targeted neonates admitted to NBU with a provisional diagnosis for sepsis between September 2017 to July 2018. Venous blood (1ml) was collected aseptically by trained clinicians and transferred into Beckton and Dickinson (BD) pediatric blood culture bottles. Culture and sensitivity assays were performed using BACTEC blood culture test protocol throughout the entire study. Maternal and neonatal demographic and clinical characteristics data were collected from the existing medical records. The study received ethical approval from the Institutional Research and Ethics committee (IREC) of Moi University and MTRH. A parental informed consent was obtained from all the mothers by a trained research assistant who explained the scope, objectives, methods, risk and benefits associated with the study. The participate data were stored in locked cabinets and databases to ensure their privacy and confidentiality. The bacterial isolates obtained were classified as either Gram positive or Gram negative. The sensitivity patterns of the various antimicrobials were assessed for the various positive isolates. Descriptive statistical techniques were used to summarize the findings as frequencies, mean and median with corresponding proportions, standard deviations and inter quartile ranges. Inferential statistics techniques such as one sample t-test and bivariate analysis were conducted to determine any significant differences and odds ratios between the predictor and outcome variables.

## RESULTS

The study enrolled 141 neonates majority of whom (57.4%) were female. The median gestational age at the time of birth was 37 (IQR: 22-45) weeks. Spontaneous vertex delivery (SVD) was the commonest among mode of delivery for 110 (78%) of the neonates enrolled, who had a median birth weight of 2400 grams (IQR: 800 - 4700) with 47.5% having a normal birth weight (Table 1).

**Table 1:** Characteristics of the neonates with neonatal sepsis at MTRH NBU. The median age of mothers enrolled was 24 (IQR: 15 - 40) years majority (54.1%; n=72) of whom were primigravida. Nearly all (93.1%; n=108) respondents attended antenatal clinic, more than half (56.6%; n=69) were unemployed with the more than one-quarter having attended a tertiary educational institution (Table 2).

| Characteristic | N | Frequency (%) / Median (IQR) |
| --- | --- | --- |
| **Gender** | 141 |  |
| Male |  | 60 (42.6%) |
| Female |  | 81 (57.4%) |
| **Gestational Age** | 125 | 37 (22 - 45) weeks |
| **Mode of Delivery** | 141 |  |
| SVD |  | 110 (78%) |
| C/S |  | 31 (22%) |
| **Birth Weight** | 139 | 2400 (800 - 4700) grams |
| ELBW |  | 6 (4.3%) |
| VLBW |  | 24 (17.3%) |
| LBW |  | 41 (29.5%) |
| Normal |  | 66 (47.5%) |
| Macrosomia |  | 2 (1.4%) |
| **Prematurity** | 141 |  |
| Preterm |  | 98 (69.5%) |
| Term |  | 43 (30.5%) |

**Table 1:** Characteristics of participants’ Mothers. Out of the 151 bacterial isolates identified at the neonatal unit of MTRH, majority (46.4%; n=70) were Klebsiella *spp* followed by *Coagulate negative staphylococcus spp* (27.8%; n=42). There were low frequencies of *Acinetobacter spp*. (6.6%; n=10), *Staphylococcus aureus* (4.7%; n=7), *Enterococcus fecalis* (3.3%; n=5), *Escherichia coli* (2.6%; n=4) reported (Table 3)

| Characteristic | N | Frequency (%) / Median (IQR) |
| --- | --- | --- |
| **Age** | 136 | 24 (15 - 40) years |
| **Parity** | 133 |  |
| Primigravida |  | 72 (54.1%) |
| Multigravida |  | 61 (45.9%) |
| 2 |  | 29 (21.8%) |
| 3 |  | 20 (15%) |
| 4 |  | 2 (1.5%) |
| 5 |  | 4 (3%) |
| 6 |  | 3 (2.3%) |
| 7 |  | 3 (2.3%) |
| **ANC Attendance** | 116 |  |
| Yes |  | 108 (93.1%) |
| No |  | 8 (6.9%) |
| **UTI** | 130 |  |
| Yes |  | 19 (14.6%) |
| No |  | 111 (85.4%) |
| **Employment Status** | 122 |  |
| Employed |  | 53 (43.4%) |
| Unemployed |  | 69 (56.6%) |
| **Level of Education** | 132 |  |
| Primary |  | 50 (37.9%) |
| Secondary |  | 45 (34.1%) |
| Tertiary |  | 37 (28%) |

**Table 2:** Frequency of Bacterial Isolates at MTRH Neonatal Unit. *Klebsiella spp*. (a gram-negative bacteria) was sensitive to meropenem (OR=3.298; 95% CI: 2.219-4.902), amikacin (OR=1.116; 0.920-1.354) and cefepime (OR=1.157; 0.167-8.002). However, there were significantly higher odds of its resistance to Vancomycin (OR=2.455; 1.888-3.192, p<0.001) as shown on Table 3.

| BACTERIAL ISOLATE | Gram stain | n | Proportion (%) |
| --- | --- | --- | --- |
| *Klebsiella spp.* | Negative | 70 | 46.4 |
| *Coagulate negative staphylococci* (CoNS) | Positive | 42 | 27.8 |
| *Acinetobacter baumannii.* | Negative | 10 | 6.6 |
| *Staphylococcus aureus spp* | Positive | 7 | 4.7 |
| *Enterococcus faecalis* | Negative | 5 | 3.3 |
| *Enterobactor spp* | Negative | 4 | 2.6 |
| *Escherichia coli* | Negative | 4 | 2.6 |
| *Methicillin susceptible staphylococcus aureus* (MSSA) | Positive | 3 | 2 |
| *Alcaligenes faecalis.* | Negative | 1 | 0.7 |
| *Methicillin resistant staphylococcus aureus* (MRSA) | Positive | 1 | 0.7 |
| *Micrococcus spp.* | Negative | 1 | 0.7 |
| *Pseudomonas auroginosa* | Negative | 1 | 0.7 |
| *Streptococcus viridans* | Positive | 1 | 0.7 |
| **TOTAL** |  | **151** | **100** |

**Table 3:** Klebsiella spp Antibiotic Sensitivity and Resistance Patterns. Gram positive bacteria (CoNS) was sensitive to vancomycin (OR=5.710; 3.478-9.374) and amikacin (OR=1.497; 0.884-2.535), but resistant to the rest. There was a statistically significant association between CoNS and resistance to meropenem (p<0.001) as shown on Table 4

| Antibiotic | Pearson chi-square test |  |  |  |  | p-value |
| --- | --- | --- | --- | --- | --- | --- |
|  |  | *Klebsiella spp.* | Odds Ratio | 95% Confidence Interval of the Difference |  |  |
|  | n(%) |  |  | Lower | Upper |  |
| Vancomycin | 70 (100) | Resistant | 2.455 | 1.888 | 3.192 | <0.001 |
| Meropenem | 57 (81.4) | Sensitive | 3.298 | 2.219 | 4.902 | <0.001 |
| Ceftriaxone | 66 (94.3) | Resistant | 1.076 | 0.990 | 1.061 | 0.161 |
| Amikacin | 54 (77.1) | Sensitive | 1.116 | 0.920 | 1.354 | 0.270 |
| Cefepime | 68 (97.1) | Sensitive | 1.157 | 0.167 | 8.002 | 0.882 |
| Gentamycin | 66 (94.3) | Resistant | 1.076 | 0.990 | 1.061 | 0.161 |
| Cefotaxime | 67 (95.7) | Resistant | 1.077 | 0.983 | 1.180 | 0.122 |

**Table 4:** Coagulase Negative Staphylococcus (CoNS) antibiotic sensitivity and resistance. Neonatal risk factors such as spontaneous vaginal delivery (p-value = 0.806), hospital delivery (0.559), prematurity (0.297), Low birth weight (0.461) and 5-minute APGAR score of ≤6 (0.287) were not associated with neonatal sepsis (Table 5).

| Antibiotic | Pearson chi-square test |  |  |  |  |  |
| --- | --- | --- | --- | --- | --- | --- |
|  | n(%) | CoNS | Odds Ratio | 95% Confidence Interval |  | p-value |
|  |  |  |  | Lower | Upper |  |
| Cefepime | 42 (100) | Resistant | 1.038 | 1.001 | 1.077 | 0.208 |
| Cefotaxime | 41 (97.6) | Resistant | 1.086 | 1.004 | 1.175 | 0.116 |
| Vancomycin | **33 (78.6)** | **Sensitive** | 5.710 | 3.478 | 9.374 | **<0.001** |
| Penicillin | 31 (73.8) | Resistant | 0.985 | 0.937 | 1.036 | 0.481 |
| Meropenem | 38 (90.5) | Resistant | 2.739 | 2.061 | 3.642 | **<0.001** |
| Amikacin | **27 (64.3)** | **Sensitive** | 1.497 | 0.884 | 2.535 | 0.142 |
| Gentamycin | 36 (85.7) | Resistant | 1.005 | 0.868 | 1.162 | 0.951 |
| Ceftriaxone | 40 (95.2) | Resistant | 1.070 | 0.974 | 1.176 | 0.238 |

**Table 5:**
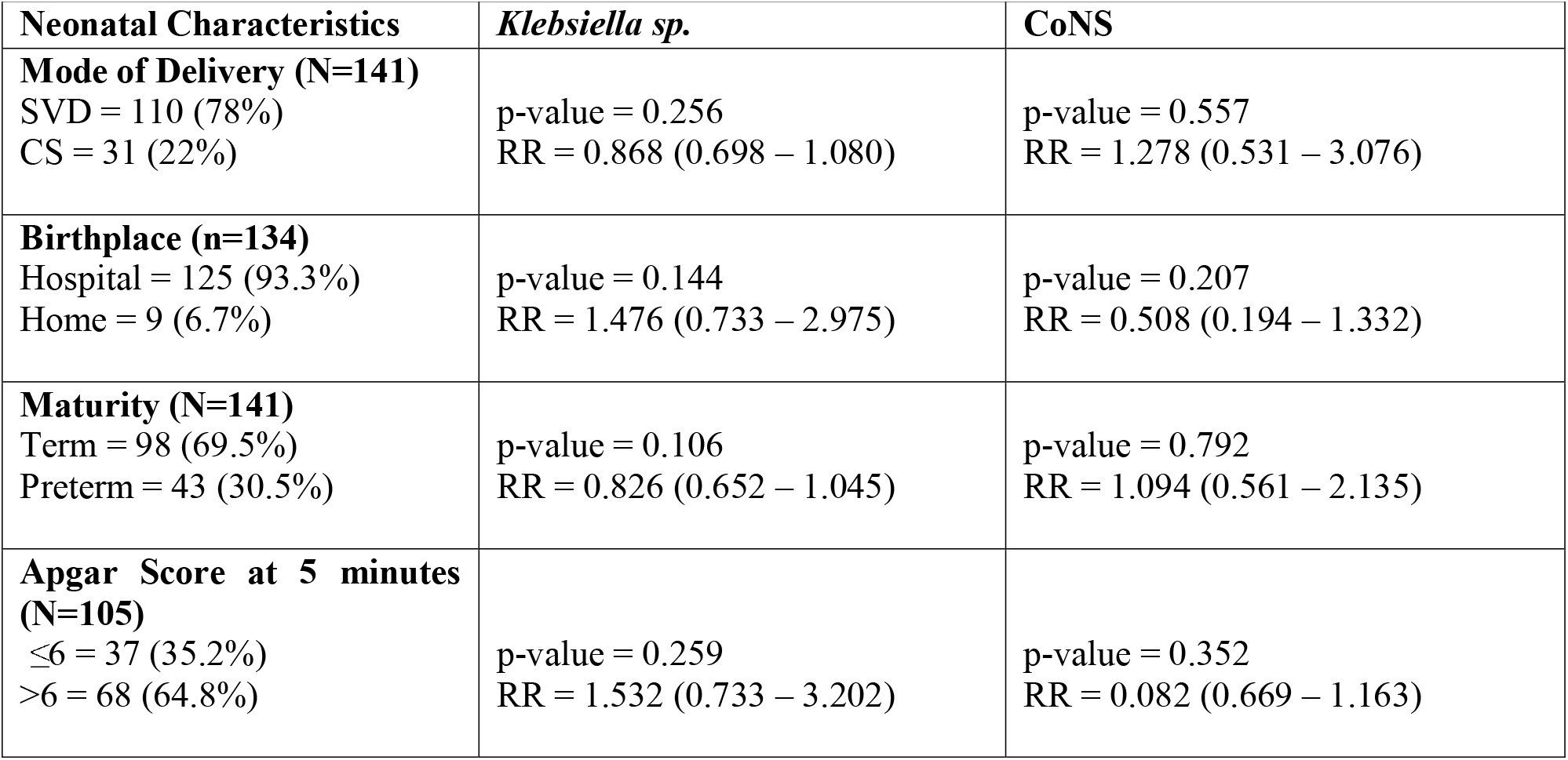
Neonatal Risk factors associated with Neonatal Sepsis. There were no statistically significant relationships demonstrated between maternal characteristics and occurrence of neonatal sepsis. Primiparity (p-value = 0.735), Intrapartum pyrexia (0.395), maternal age (0.397), level of education (0.426), urinary tract infections (0.918), prolonged rapture of membranes and mode of delivery (0.808) as shown on Table 7.

**Table 6:** Maternal Risk Factors Associated with Neonatal Sepsis.

| Maternal Characteristics | All Infections (p-value) | *Klebsiella sp.* | CoNS |
| --- | --- | --- | --- |
| **Parity**<br>(Primi/ Multigravida) | 0.783 | p-value = 0.322<br>RR – 1.116 (0.895 – 1.391) | p-value = 0.484<br>RR – 0.787 (0.401 – 1.543) |
| **Intrapartum Pyrexia** | 0.395 | p-value = 0.814 | p-value = 0.331 |
| **Maternal Age** | 0.327 | p-value = 0.630 | p-value = 0.404 |
| **Level of Education** | 0.426 | p-value = 0.365 | p-value = 0.191 |
| **UTI in pregnancy** | 0.918 | p-value = 0.304 | p-value = 0.164 |

## DISCUSSION

Neonatal sepsis is caused by various pathogenic bacteria that can be classified as either gram positive or gram negative based on their gram staining patterns. In this study, the leading causative organism of neonatal sepsis among all the enrolled children was *Klebsiella spp* (gram-negative bacteria) accounting for nearly half of all the bacteria isolated. The second most common bacteria isolated among the neonates enrolled in the study was CoNS (gram-positive bacteria) that accounted for more than one-quarter of all the bacteria isolated. Although there were other bacteria isolated from the neonates enrolled, their proportions were negligible with *Acinetobacter baumannii* (a gram-negative bacteria) being the third most prevalent bacteria isolated.

These results are consistent with those from previous studies conducted in Kenya [19] and India [20]. In the Kenyan study conducted at Kilifi District Hospital, *Klebsiella spp* was the most prevalent of all the bacterial isolates and the leading gram negative bacteria seen [19]. This similarity could be attributed to the fact that both studies were conducted in public hospitals in the same country where infection control strategies and study population are similar.

However, bacterial isolates proportions differ from those reported in Kenya [21], Bangladesh [22] and Arab States in the Gulf region – Kuwait, Saudi Arabia and the United Arab Emirates - [23]. In the Kenyan ten year (2000 - 2009) retrospective review conducted in Aga Khan university hospital – a private teaching hospital -, CoNS was the most prevalent (27%) of the bacteria isolated from the neonates followed by *Klebsiella spp* [21]. This difference could be attributed to difference in study settings and design.

In the Bangladesh study conducted at Ad-din Medical College Hospital (AMCH) in Dhaka; it was reported that CoNS (68.4%) was the most prevalent followed by *Acinetobacter baumannii* at 18.4% [22]; however, there was no *Klebsiella spp* reported. This difference could be attributed to difference in study designs as the Bangladesh study was prospective over a period of nine-months while the current study was cross-sectional.

In the Gulf Region states [23], it was reported that CoNS was the most prevalent (34.65%) followed by *Klebsiella spp (*22.8%*), E. coli* (4.845) and *Acinetobacter spp* (4.59%). The prospective study conducted in the NICUs of Kuwait, United Arab Emirates (UAE) and Saudi Arabia was different from the current study due to its study design, target population (late onset sepsis) and sample size (n=780); further explaining the difference in study findings.

*Klebsiella spp* was sensitive to meropenem (OR=3.298; 95% CI: 2.219-4.902), amikacin (OR=1.116; 0.920-1.354) and cefepime (OR=1.157; 0.167-8.002). However, there were significantly higher odds of its resistance to Vancomycin (OR=2.455; 1.888-3.192, p<0.001) and Gentamycin (OR=1.907; 1.86-1.95, p=0.163). The sensitivity data in this study were similar to those found in India [20] found Klebsiella to be sensitive to amikacin and resistant to penicillin. Gentamicin sensitivity results are also similar to a study in the Gulf Region States [24] which reported Klebsiella resistance to Gentamicin (8%). The findings of the current study contrast those from a Kenyan study which found Klebsiella to be sensitive to Gentamicin (72.4%); however, in the Aga Khan hospital study, sensitivity of Klebsiella to amikacin were similar at 94.1% [21].

Coagulase negative Staphylococcus aureus (CoNS) was sensitive to vancomycin (OR=5.710; 3.478-9.374) and penicillin (OR=2.595; 0.166-40.550), but resistant to the rest. These findings are similar to an Indian study which found CoNS to be resistant to Gentamicin, ceftriaxone and Ceftazidime. However, CoNS was sensitive to vancomycin and penicillin. The findings match those from Bangladesh [22] who found CoNS to be sensitive to Vancomycin (74%) while resistant to Meropenem (22%) and Amikacin (0%). In a previous Kenyan study [21], it was reported that CoNS was sensitive (88.9%) to amikacin and Gentamicin (83.4%) which contrast the findings of the current study. This could be attributed to the abuse of gentamycin (a first line antimicrobial) over the years without laboratory diagnostic confirmation.

In this study, there was no statistically significant relationship demonstrated between maternal characteristics and occurrence of neonatal sepsis. Primiparity (p-value = 0.735), Intrapartum pyrexia (0.395), maternal age (0.397), level of education (0.426), urinary tract infections (0.918), prolonged rapture of membranes and mode of delivery (0.808). These findings are similar to studies conducted in Saudi Arabia [25], Uganda [26] and South Korea [27]. In a study conducted at King Abdul Aziz Specialist Hospital in Taif, Saudi Arabia [25], it was reported that there was no statistically significant association between intrapartum pyrexia (0.110) and prolonged rapture of membrane (0.210). Although both studies adopted a cross-sectional study design, however, in Saudi Arabia, the neonates were stratified into three groups (proven early-onset neonatal sepsis, clinical early-onset neonatal and negative infectious status) while the current study only had a single group of neonates with sepsis. At Uganda’s Mulago hospital [26], the study assessed the aetiology and risk factors for neonatal sepsis and determined that primiparity (0.23), intrapartum pyrexia (0.060), prolonged rapture of membranes (0.140) and mode of delivery (0.070) was not significantly associated with the occurrence of neonatal sepsis. These findings were also similar to those from South Korea [27] where no statistically significant association between prolonged rapture of membranes (0.840) and occurrence of neonatal sepsis was reported. In contrast, a study conducted in Ethiopia’s public hospitals of Mekelle City [17] reported that the occurrence of neonatal sepsis was significantly associated with intrapartum pyrexia (AOR = 6.08), urinary tract infections (AOR = 5.23) and prolonged rapture of membranes (AOR = 7.4).

Furthermore, the selected neonatal risk factors such as spontaneous vaginal delivery, hospital delivery, prematurity, low birthweight and a 5-minute APGAR score of ≤6 were not found to be significantly associated with neonatal sepsis. This was also the case with the findings reported in Saudi Arabia [25], Uganda [26] and South Korea [27].

## CONCLUSIONS AND RECOMMENDATIONS

This study determined that the main bacterial causes of neonatal sepsis at a teaching hospital in Kenya were *Klebsiella spp*. and CoNS. Both the gram positive and gram-negative bacteria had good sensitivity to meropenem and amikacin. The risk factors evaluated were not associated with the occurrence of neonatal sepsis. Klebsiella being one of the known nosocomial infections, improvement in infection control in the unit is recommended. There is need for evidence-based review of empirical antibiotic therapy regimen containing penicillin, gentamycin, and ceftriaxone due to the prevailing high resistance levels. Future studies targeting specific risk factors to be conducted to validate this study’s finding.

### What is already known on this topic

Neonatal sepsis is one of the leading causes of morbidity and mortality among newborns globally [28]. Its prevalence in higher in developing countries compared to the developed ones with an estimated prevalence of 12% and 28% in Nigeria [5] and Kenya [6] respectively. A combination of neonatal, maternal and environment risk factors could predispose neonates to sepsis. Most of the neonates presenting with sepsis are likely to succumb to the disease if it is not promptly addressed [2].

### What this study adds

Because the major sepsis causative organisms in the neonatal units of major referral facilities in Kenya is not well documented, there is need to determine these causative organisms to inform localized infection control strategies, improve on patient care and clinical outcomes. This study reports neonatal sepsis causative organisms, their antibiotic sensitivity patterns and risk factors associated with neonatal sepsis at a resource limited neonatal unit in Kenya. The findings will inform proper antibiotic stewardship to help optimize antimicrobial therapy, ensure improved clinical outcomes and lowering the risk of subsequent antibiotic resistance.

## Data Availability

All the data supporting this study will be made available.

## Acknowledgements

The authors would like to thank the parents of the neonates who participated in this study, the hospital management at Moi Teaching and Referral Hospital, the academic faculty and students at the Department of Child Health and Paediatrics in the School of Medicine at Moi University – Eldoret, Kenya.

## Competing Interests

The authors declare no competing interests.

## Author Contribution

1. **BMA, JS and WN:** Substantial contributions to conception and design of the study.
2. **BMA, JS and WN:** Acquisition of data, analysis and interpretation of data designed and carried out data collection and participated in drafting the manuscript.
3. **BMA, JS and WN:** drafting the article or revising it critically for important intellectual content.
4. **BMA, JS and WN:** Final approval of the version to be published intellectual content. They also gave the final approval of the version to be published and have agreed to be accountable for all aspects of this work.

## Notes

### Competing Interest Statement

The authors have declared no competing interest.

### Funding Statement

This study did not receive any external funding.

### Author Declarations

Institutional Research and Ethics Committee of Moi University and Moi Teaching and Referral Hospital

